# Developing an initial program theory for peer support groups for substance-affected family members: a participatory realist evaluation

**DOI:** 10.64898/2025.11.28.25341232

**Authors:** Jennifer Hawkins, Anne Whittaker, Amy Salmon, Frances Kenny, Morgan R. King-Roskamp

## Abstract

**Background:** Amidst an unregulated drug crisis in Canada, the needs of affected family members have been largely ignored. Evidence supports the efficacy of peer support group interventions in health and social care; however, a better understanding of *how* peer support groups produce outcomes is required for successfully implementing interventions for substance-affected family members.

**Methods:** The study uses a participatory realist evaluation framework to develop an initial program theory (IPT). Data collection includes interviews of program staff and participants, program evaluation surveys, internal program documents, and observational field notes. We employ deductive and inductive coding and retroductive analysis theorizing contexts, mechanisms, and outcomes configurations (CMOCs).

**Results:** We present three nested implementation contexts containing 10 CMOCs. Our IPT suggests: Strong organizational support in an environment that values lived expertise promotes program sustainability; a skilled facilitator with lived experience will leverage unique approaches to foster a positive group culture; a positive group culture activates the program’s pillars of emotional support, education and skill-building to produce reduced social isolation, improved emotional wellbeing, increased self-efficacy, and improvements in relationships.

**Conclusion:** This participatory realist evaluation presents an IPT of a peer support group for substance-affected families that will inform future implementation and evaluation of family supports.

## Introduction

### Background

In 2016, the Provincial Health Officer in British Columbia (BC), Canada, declared a public health emergency in response to a rising number of deaths from unregulated drugs contaminated with fentanyl and fentanyl-analogues. In the 10 years since, the crisis shows little sign of abating: toxic drug poisonings have become the leading cause of death for British Columbians aged 10-59 (BC Coroner’s Service, 2023). Despite a concerted effort to apply targeted interventions and policy changes, the needs of affected family members have been largely ignored (Hawkins, Salmon, et al., 2025; Mathias, Duff, et al., 2025). These social connections can constitute critical means of support, and affected family members need and deserve support in their own right (Fernando et al., 2022; Mathias, Auger, et al., 2025; Orford et al., 2013; Rokiyah et al., 2024; Selbekk et al., 2018). Furthermore, research indicates that unsupported carers represent a considerable healthcare burden (Denomme & Benhanoh, 2017; Orford et al., 2013; Slaunwhite et al., 2017; Soares et al., 2016), and individualistic responses to addiction that ignore affected family members are failing to act on available evidence (Orford et al., 2013; Selbekk & Sagvaag, 2016).

Our community-academic research partnership in the Fraser East of BC has worked closely with people with lived experience of illicit substance use as well as affected family members and community service providers (Hawkins, Kniseley, et al., 2025). In two linked studies, we explored reasons why people use alone, as well as affected family members’ experiences during the toxic drug crisis. The quality and nature of personal relationships emerged as a particularly salient factor influencing people’s complex decisions about drug use (Fernando et al., 2022), and both studies indicated that isolation could heighten overdose-related risks when family members have exhausted their ability to cope (Fernando et al., 2022; Hawkins, Salmon, et al., 2025). Family members also reported exacerbation of stress from the toxicity of the drug supply, experiencing the possibility of “getting the call” about a loved one’s death as an ever-present reality. The intensity of this stress included a preponderance of negative interactions with formal systems of care as well as an overall paucity of social support; participants were often dealing with what they perceived as “failures” of the substance-use care systems as well as a lack of understanding within their informal support networks (Fernando et al., 2025). Despite this overall dearth of support, study participants observed that peer support offered the potential for a significant alleviation of acute stress burdens (Fernando et al., 2025; Hawkins, Salmon, et al., 2025).

Peer support (also called ‘mutual aid’) can offer a uniquely effective form of social capital (Natung et al., 2025). Orford and colleagues report that, for various reasons, substance-affected individuals’ primary social networks can be unsupportive and unhelpful (Orford, Velleman, et al., 2010). This was also reflected in our findings, where intra-family conflict often increased experiences of stress; however, stress was significantly mitigated by social support from peers with shared lived experience (Fernando et al., 2025; Hawkins, Salmon, et al., 2025). The literature widely supports the efficacy of peer-support interventions. Numerous systemic reviews and meta-analyses of peer support in mental health settings report positive outcomes related to hope, empowerment, quality of life, reduced symptoms, recovery, and reduced social isolation and hospitalization (Bellamy et al., 2017; Kundurthi et al., 2025; Pfeiffer et al., 2011; Shorey & Chua, 2022; White et al., 2020). Systematic reviews also provide evidence for improvement in psychological health, social support, self-efficacy and clinical outcomes (Berg et al., 2021; Haines et al., 2018). While reviews can be hampered by heterogeneity in intervention design, implementation, and the lack of distinction between peer support groups and one-on-one peer support, a recent evidence map of high-quality studies for peer support in the domains of health and social care found strong evidence for the efficacy of peer support in providing educational support, help with self-care and self-management, and social support (Price et al., 2022). Family members are increasingly viewed as critical stakeholders in the toxic drug response (Mathias, Auger, et al., 2025), and recent studies provide evidence of positive outcomes from peer support, such as increased self-efficacy and reduced stress (Kelly et al., 2017; Peart et al., 2023, 2024).

### Purpose

While evidence shows peer support group participation is linked to improved outcomes for affected family members, the literature contains little information on *how* peer support groups produce these outcomes (Garn et al., 2021). In recent years, realist research has emerged as a theory-driven paradigm that addresses the complexities inherent in socially contingent public health programming (Jagosh, 2019). A key marker of quality in realist studies is ensuring rigour in the development of an initial program theory (IPT) (Jagosh et al., 2022). This paper presents the first step in realist research on this topic by describing the development of an IPT for a peer support group for substance-affected family members. In so doing, our goal is to support broader implementation and realist evaluation of peer-based support for substance-affected family members.

## Methods

Our realist approach was embedded in a broader community-based participatory research (CBPR) study examining interventions for substance-affected family members (Hawkins, Kniseley, et al., 2025). A working group of the CBPR study conducted this research in partnership with substance-affected family members, including staff from the program under study as well as peer consultants supporting our research team. We present our work using the RAMESES II Quality Standards for Realist Evaluation (G. Wong et al., 2016). Table 1 contains the checklist of standards. Ethical approval for the research was granted by the University of British Columbia-Providence Health Care Research Ethics Board (H23-03688). All participants gave informed consent.

**Table 1.**
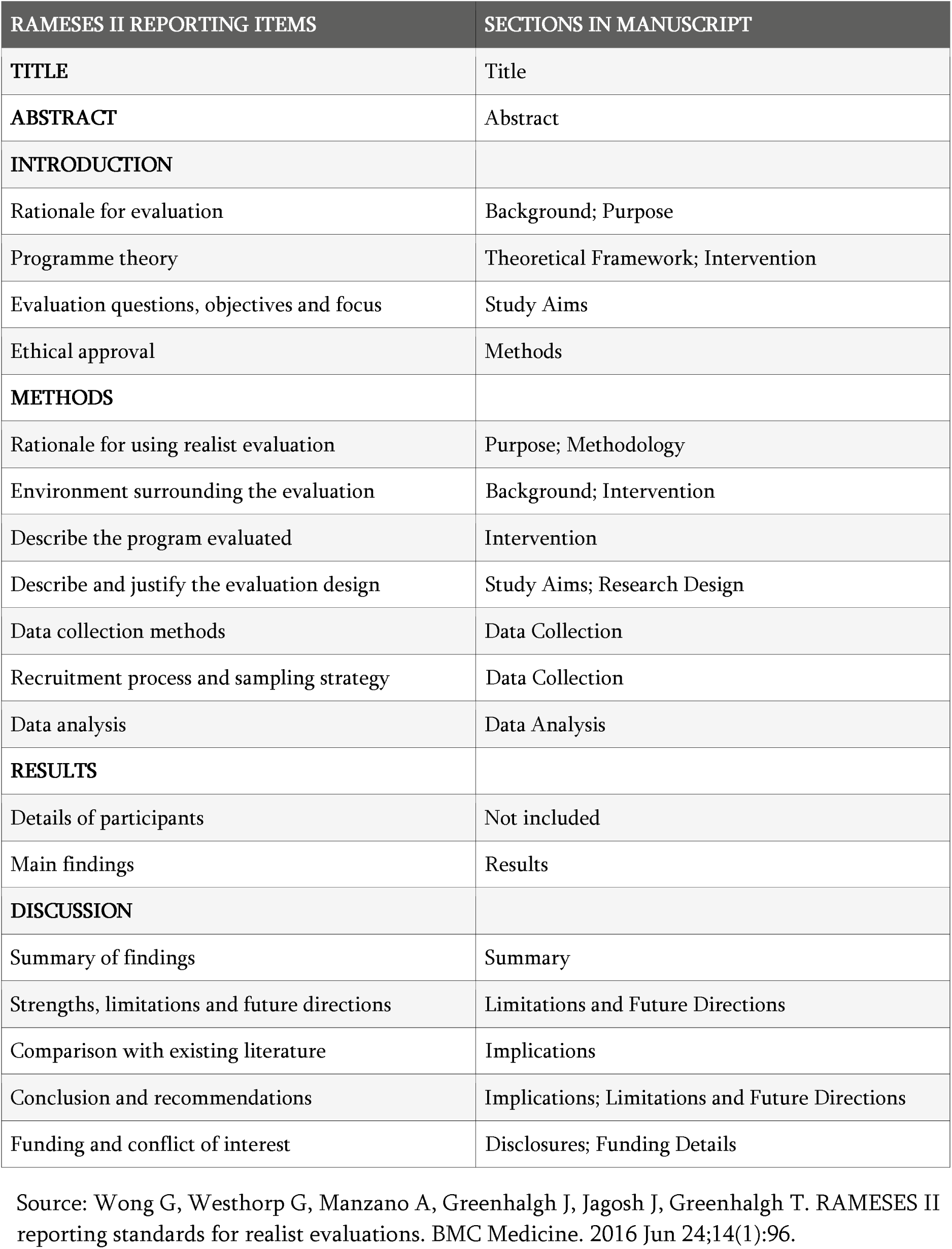
Document sections for RAMESES II reporting criteria.

| RAMESES II REPORTING ITEMS | SECTIONS IN MANUSCRIPT |
| --- | --- |
| **TITLE** | Title |
| **ABSTRACT** | Abstract |
| **INTRODUCTION** |  |
| Rationale for evaluation | Background; Purpose |
| Programme theory | Theoretical Framework; Intervention |
| Evaluation questions, objectives and focus | Study Aims |
| Ethical approval | Methods |
| **METHODS** |  |
| Rationale for using realist evaluation | Purpose; Methodology |
| Environment surrounding the evaluation | Background; Intervention |
| Describe the program evaluated | Intervention |
| Describe and justify the evaluation design | Study Aims; Research Design |
| Data collection methods | Data Collection |
| Recruitment process and sampling strategy | Data Collection |
| Data analysis | Data Analysis |
| **RESULTS** |  |
| Details of participants | Not included |
| Main findings | Results |
| **DISCUSSION** |  |
| Summary of findings | Summary |
| Strengths, limitations and future directions | Limitations and Future Directions |
| Comparison with existing literature | Implications |
| Conclusion and recommendations | Implications; Limitations and Future Directions |
| Funding and conflict of interest | Disclosures; Funding Details |

Source: Wong G, Westhorp G, Manzano A, Greenhalgh J, Jagosh J, Greenhalgh T. RAMESES II reporting standards for realist evaluations. BMC Medicine. 2016 Jun 24;14(1):96.

### Methodology

Realist evaluation offers a theory-based approach that examines how interventions work in dynamic contexts that can defy both the logic of counterfactual trials and formulaic replication (Flint-Taylor et al., 2023; Jagosh, 2019). It was developed by Pawson and Tilley to account for the complexity of real life contexts that impact outcomes, examining not just *whether or not* an intervention works, but *what* works, *for whom*, *under what circumstances, how,* and *why* (Pawson & Tilley, 1997). Realist paradigms have increasingly contributed to implementation science by addressing the challenges inherent in replication (Sarkies et al., 2022). Realist evaluation’s main principle maintains that all interventions contain underlying theories that explain the relationship between complex contexts, the mechanisms they activate, and the resulting outcomes (Garn et al., 2021). Its primary analytical tool is the Context-Mechanism-Outcome configuration (CMOC), whereby program theories provide generative explanations about how or why the program works that are then tested in real-world settings (Jagosh, 2019). Table 2 contains definitions of CMO categories (Dopson & Fitzgerald, 2007; Jagosh, 2019; Jagosh et al., 2015; Pawson & Tilley, 1997).

**Table 2.** CMO heuristic definitions.

| CONTEXT-MECHANISM-OUTCOME (CMO) HEURISTIC |
| --- |
| **CONTEXT:** Context can be defined as the “backdrop” to implementation. It includes all factors that are not part of the program or intervention itself but have an impact on outcome, such as demographics, legislation, or cultural norms (Jagosh, 2019). |
| **MECHANISM:** Mechanisms are the combination of resources (intended and unintended) created by a program and the response to those resources (cognitive, emotional, motivational reasoning, etc.) by stakeholders (Pawson & Tilley, 1997). Mechanisms can pertain to why participants choose (or choose not) to participate in interventions or internalize health knowledge or behavior change from the intervention (Jagosh et al, 2015). Mechanisms will only activate in the right contexts. |
| **OUTCOMES:** Outcomes are a result of a program activating multiple mechanisms. Because mechanisms have different effects on different subjects in different situations, an intervention can produce multiple outcomes. Realist evaluations thus examine outcome patterns in a theory testing approach (Pawson and Tilly, 1997; Jagosh, 2019). |
| **CMOCs:** CMO configuring is a heuristic used to generate causative explanations about outcomes in the observed data. A CMO configuration may be about the whole program or only to certain aspects. One CMO may be embedded in another or configured in a series (ripple effect in which the outcome of one CMO becomes the context for the next in the chain of implementation steps). Configuring CMOs is a basis for generating and/or refining the theory that becomes the final product (Jagosh et al., 2015). |

Our realist evaluation of a peer support program model also employed participatory approaches. Participatory research methods have become increasingly prevalent in the design and implementation of health interventions (Belaid et al., 2023). As a transformative paradigm that contextualizes processes and results to local settings and groups, participatory approaches expand the capacity of translational sciences to disseminate effective interventions and increase health equity (Belaid et al., 2023; Jagosh et al., 2012; Wallerstein & Duran, 2010). In CBPR, community members are involved as equal partners at all stages in research that requires respect as a cornerstone (J. Wong et al., 2025). When combined with realist research, participatory approaches unite the causal insights of theory with the insights of lived experience (Halsall et al., 2022).

### Theoretical Framework

Our prior research findings aligned closely with the stress-strain-coping-support (SSCS) model (Fernando et al., 2025), a theoretical framework resulting from more than two decades of qualitative research with affected family members. The SSCS model posits that caring for a loved one in addiction is a chronically stressful experience leading to strain that frequently manifests as ill-health (Orford, Copello, et al., 2010; Orford et al., 2013; Orford, Velleman, et al., 2010). According to the SSCS model, both formal and informal social support that is informational, emotional, behavioural and material can effectively mediate against the cumulative stress burden of substance-affected family members, improving their ability to cope and thus their overall wellbeing (Orford, Copello, et al., 2010). The SSCS model is one of the few frameworks that is non-pathologizing towards family members and affirms their need for support in their own right (Fernando et al., 2025; Peart et al., 2024). When we began to engage interested community members with these findings, we encountered a peer support group intervention, Parents Forever (PF), that aligned closely with the SSCS model in its philosophy and methods (Salmon et al., 2025).

### Intervention

Parents Forever offers mutual support for affected family members through one-to-one intake, two-hour weekly support groups, regular educational sessions, connection to resources, and, when needed, one-to-one support provided by a facilitator with lived experience. The program defines “mutual support” as support that family members both give and receive; mutual support differs from therapy or counselling, and it discourages advice-giving. Instead, mutual support acknowledges the hard-earned expertise and wisdom of lived experience, encouraging participants to help each other find strategies to cope in an environment of compassion and understanding.

The program was founded by Frances Kenny, a parent with lived experience as an affected family member who coordinated the program from 2000-2025. PF was based on Kenny’s experiences with the Parents Together program of the Boys and Girls Club (BGC) of Canada and was supported by BGC during its inception and early years. Starting in 2015, PF was financially and administratively supported through public health authority funding that was contracted through a non-profit entity; however, when we first encountered the program, its future was uncertain due to Kenny’s approaching retirement. Additionally, despite its clear alignment with the SSCS model, it was unclear whether its locally-touted success was solely due to the skills, personality and dedication of its founder, or whether its program model was both responsible for reported positive results and broadly replicable.

### Study Aims

Our realist approach focused on the following questions: How do PF staff and program participants describe the impacts of the program? What are the qualities and approaches of PF’s coordinator and service delivery model, and how do they contribute towards program outcomes? How have the contexts surrounding PF enabled the longevity and efficacy of the program? We aimed to develop an IPT for a peer support group model that could be more broadly implemented and evaluated using a realist approach.

### Research Design

The study was primarily designed and conducted by members of a working group of our broader CBPR team. First, we used the Social Ecological Model (SEM) to map contextual factors unique to the toxic drug emergency and local environment based on our prior findings and the lived experience of team members (Fernando et al., 2022, 2025; Hawkins, Salmon, et al., 2025). We also consulted with a working group of non-PF peers, who had not received peer support group interventions, to discuss their needs and perspectives regarding Context-Mechanism-Outcome (CMO) categories. We used the map and notes from these meetings in the second phase of our data analysis described below.

To optimize respectful engagement with PF, we spent one year in regular, in-person meetings with Kenny, learning about the program’s origins, development, values, key components, mode of delivery, and hearing stories of successes and challenges. We also consulted several times with PF’s current and past clinical supervisors and other professionals who had been involved with the program’s development and implementation. This allowed us to thoroughly understand PF’s history and basic architecture. We then co-developed data collection methods with Kenny.

### Data Collection

Realist evaluation encourages the use of multiple forms of data to support theorizing (Greenhalgh et al., 2015), including the use of non-empirical literature in developing an IPT (Jagosh et al., 2022). We used two different tiers of data corresponding to two different phases of our analysis. Tier 1 consisted of semi-structured interviews with two program-affiliated staff and five program participants; results from three qualitative program evaluation surveys administered in 2019, 2021, and 2024 with a total of 53 respondents; two non-empirical articles written about PF. Tier 2 consisted of program documents, including program policies and peer-developed resources used in program delivery; notes from participant observation of three two-hour support group meetings; the SEM contextual map and notes from the non-PF peer consulting sessions. For the semi-structured interviews, we recruited purposively, with information provided to PF participants by Kenny verbally at weekly meetings and via email. Table 3 contains an outline of interview and evaluation questions in CMO categories.

**Table 3.** Interview and program evaluation respondent data.

| INTERVIEW DATA |  |  |
| --- | --- | --- |
| CMO CATEGORY | TOPIC | EXAMPLE QUESTIONS |
| Context | • Social and support environments<br>• Prior information and understanding<br>• Issues specific to the toxic drug crisis | • What other services have you or do you access for support related to your loved one's substance use?<br>• Before you began attending the group, how would you describe your level of knowledge of addiction?<br>• Did you find any helpful perspectives in your social circles?<br>• How has the public health emergency influenced how you seek support? |
| Mechanisms | • Access and availability of the program<br>• Nature of services delivered<br>• Experiences with the program<br>• Resulting feelings and opinions | • Can you describe your experience attending the first group meeting?<br>• Are there any attributes of the group you feel are particularly effective?<br>• Is there anything in particular that made you want to come back? |
| Outcomes | • Perceived benefits<br>• Potential gaps or drawbacks | • Can you describe any positive changes in your life after attending the group?<br>• Does anything ever feel too difficult for the group to help you with? |
| EVALUATION DATA |  |  |
| CMO CATEGORY | TOPIC | EXAMPLE QUESTIONS |
| Context | • Social and support environments | • What are “missing pieces” of the current supports you access? |
| Mechanisms | • Services delivered<br>• Resulting feelings and opinions | • Can you suggest any changes to the current program format?<br>• What is your level of satisfaction with feeling safe, respected, and listened to?<br>• What is your level of satisfaction relating to connections with other parents? |
| Outcomes | • Perceived benefits<br>• Potential gaps or drawbacks | • Can you describe any changes in yourself and your relationship with your loved one from Parents Forever? |

### Data Analysis

Realist analysis employs the CMO configuring heuristic as its primary analytic method (Pawson & Tilley, 1997). We conducted our analysis in two main phases: 1) initial coding to identify contexts, mechanisms and outcomes; 2) iterative analysis refining CMOs and retroductive analysis identifying theoretical pathways (CMO configuring). Retroductive analysis produces generative causal insights by using deductive and inductive reasoning that includes researcher insights from multiple sources of data (Gilmore et al., 2019). In phase 1, using NVivo 14 and tier 1 data, JH coded outcomes inductively; mechanisms and contexts were coded deductively with categories derived from the SSCS model. Additional nodes and sub-nodes were categorized inductively. Mechanisms can be notoriously difficult to determine, and the differences between contexts and mechanisms can be complex and often seem unclear (Flynn et al., 2021; Marchal et al., 2012). We therefore transferred the next phase of analysis to a series of spreadsheets to further refine the CMO categories, incorporating tier 2 data to provide further depth and clarity. The spreadsheets were then refined iteratively with JH, FK, AW and AS. Figure 1 provides an example of a spreadsheet containing candidate contexts. Using memos, NVivo codes, and collective deliberation, we then conducted retroductive analysis to finalize CMO configurations (CMOCs).

**Figure 1.**
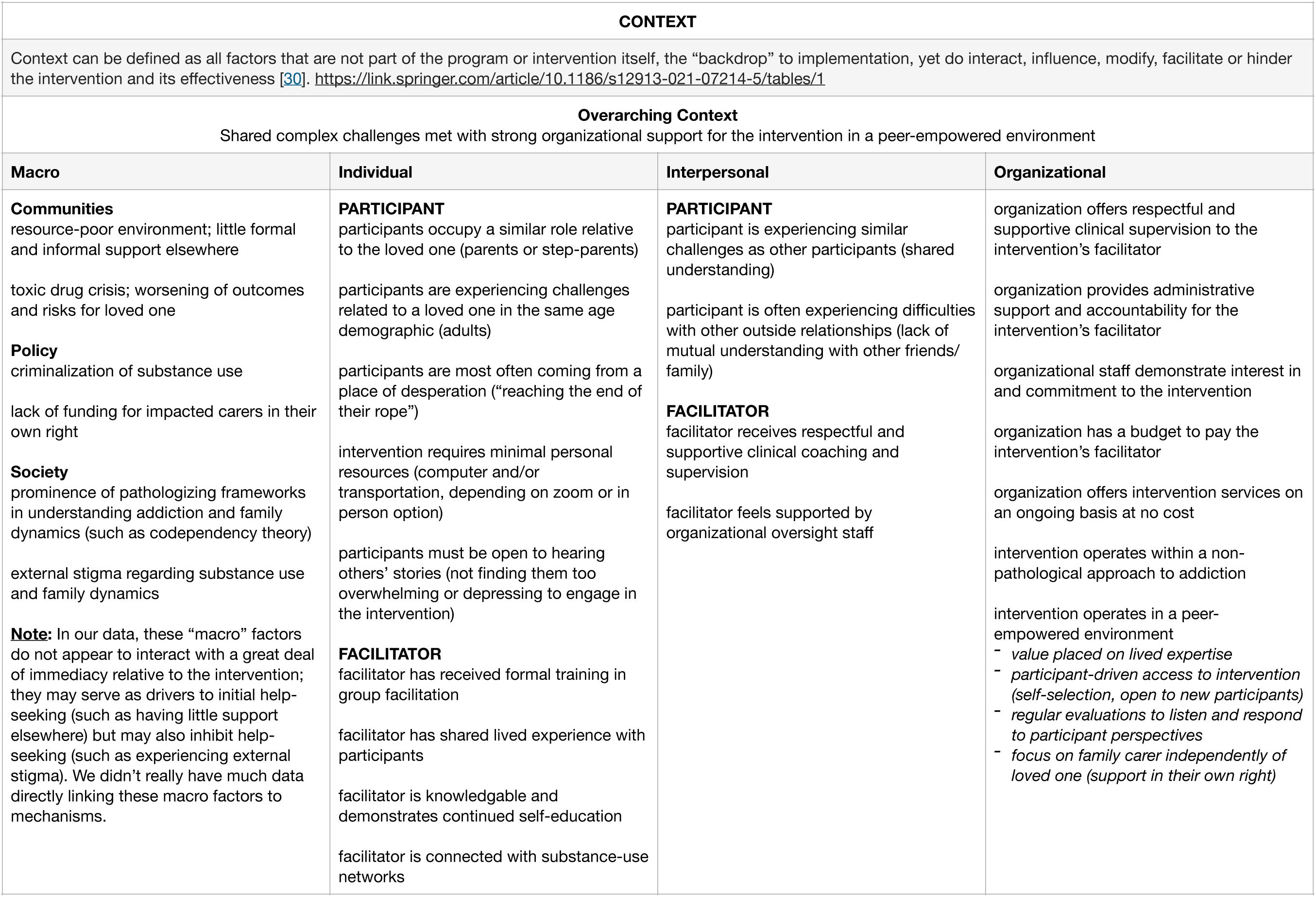
Example of spreadsheet used in phase 2 of data analysis

## Results and Findings

Our retroductive reasoning led us to apply the “ripple effect” into our CMOCs, whereby the outcome of one phase of a project becomes an aspect of context for the next phase (Jagosh et al., 2015). In this way, we arrived at a series of nested implementation contexts providing explanatory power for the program’s outcomes. Table 4 outlines 10 CMOCs embedded within three nested contexts.

**Table 4.** CMOCs in nested contexts.

| IMPLEMENTATION CONTEXT 1.0 |  |
| --- | --- |
| Strong organizational support operating in a peer-empowered environment |  |
| 1.1 | When a program receives ongoing funding (C), if the organizational operator has a commitment to the program (C) and operates in a peer-empowered environment with a focus on support for affected family members in their own right (C), the program offers ongoing services to participants at no cost (M) with an open, participant-driven access to the program (M). This results in a program that is highly accessible to potential participants (M) and supports the sustainability of the program longer-term (O). |
| 1.2 | When the program facilitator has lived experience as an affected family member (C) and demonstrates continued self-education (C), they will leverage personal understanding, empathy and expertise with training and skill development in peer support facilitation (M). This leads to the facilitator operationalizing high levels of facilitation skills (O). |
| 1.3 | If the organizational operator of the program has available clinical expertise on staff (C) that engages that facilitator respectfully (C), then the program facilitator can receive supportive clinical coaching and supervision (M), which leads to their feeling supported by organizational oversight staff (M). This support, alongside the facilitator's lived expertise (C), enables the facilitator to display high levels of personal investment, dedication and passion (O) and enhances the program's sustainability (O). |
| IMPLEMENTATION CONTEXT 2.0 |  |
| Accessible, sustainable program coordinated by a strong peer facilitator |  |
| 2.1 | When participants enter a program from a place of desperation (C), if a skilled peer facilitator (C) meets them with a relational approach (M), they will feel safe and connected to the facilitator and will develop trust in the facilitator and the group process (M). This leads to higher levels of engagement with the program (O) and contributes to a strong group culture (O) and a genuinely connected coordinator (O). |
| 2.2 | When a strong peer facilitator (C) coordinates a group that has high levels of similar lived experience (C), if the facilitator provides a non-pathological approach to addiction (M) to shape the character of the group (M), participants respond with a willingness to follow group norms and a feeling of trust and appreciation for the facilitator and the group process (M). This cultivates a strong group culture of compassion, non-judgement, acceptance and understanding (O) with high levels of engagement with the support model (O). |
| 2.3 | If the group is run by a strong peer facilitator (C) who is supported with ongoing clinical oversight (C), the coordinator maintains a balance of structure with flexibility and adaptation (M) that responds to participant feedback (M) and fosters confidence in the group process (M), which engenders trust, gratitude and appreciation for the facilitator and group (M). This creates a genuinely connected facilitator (O) and high levels of engagement with the program (O). |
| 2.4 | When participants come to the group from a place of desperation (C) and the facilitator applies a relational approach (M) with an orientation towards "reasonable hope" (M) that fosters a sense of positivity while acknowledging hardships (M), participants experience an increase in hope (M) that generates higher levels of engagement with the program (O) and a positive group culture (O). |
| IMPLEMENTATION CONTEXT 3.0 |  |
| Strong, positive, engaged group culture supporting key program pillars |  |
| 3.1 | In an external environment saturated with pathologizing frameworks for addiction (C) with very few additional support resources (C), participants will be open (M) to the program's integration and prioritization of education (M). When the facilitator also displays continued self-education (C1) and is connected with substance use networks (C1), participants experience increased knowledge and understanding (M) that leads to increased self-efficacy (O). |
| 3.2 | If participants are engaged with the program (C) and come with a willingness to hear others' stories (C), they will be open to the program's implementation of peer support (M). If the group operates within a strong culture of acceptance, empathy, non-judgement and compassion (C), the integration of structured, non-therapeutic peer support leads to participants feeling understood (M), experiencing genuine caring for other group members (M) and developing supportive relationships with other group members (M). This results in decreased social isolation (O) and improved emotional wellbeing (O). |
| 3.3 | When participants are ready to apply program resources (C) to difficulties they face in their relationships (C), and when the program operates within a strong group culture (C), participants will display openness to the interventions components (M). When the program integrates a collective, developmental model of skill-building (M) where peers are included in the development and application (M) by a connected and responsive facilitator (C), participants can experience positive changes in their communication and connection with their loved one (M), leading to more tensile connections (O), emotion regulation and decreased conflict (O). |

### Context 1: Strong organizational support operating in a peer-empowered environment

#### I guess we’ve all been in meetings where you’re just kind of passing around a bowl of chips and there’s no sort of curation of best practice. So it gives it a layer of credibility. - PF Participant

At the time of our research, PF had been operating for 25 years with a high volume of uptake and no interruption of service, including during the COVID-19 pandemic. Foundational contexts enabling this sustainability consisted of ongoing funding with organizational support that operated in a peer-empowered environment. We found that ongoing funding would not likely have been adequate on its own; organizational operators needed to appreciate the wisdom of lived experience and view support as inherently valuable regardless of whether family members’ loved one’s engaged in substance use services. This reflects the SSCS model’s assertion that affected family members require support “in their own right” (Copello et al., 2010). With ongoing organizational support that valued lived expertise, PF was able to offer participant-driven access to its services that ultimately proved highly sustainable.

Peer-empowered organizational values and ongoing funding also provided paid support for a program facilitator (Kenny) with lived experience who did not hold the professional counselling credentials often required to run support groups. However, Kenny was able to receive training and also exhibited passion and curiosity for ongoing education on related topics and resources. One respondent’s perspective was reflected throughout our data: “It’s so important that the facilitator has a vast background of education, contacts, and connections, and—as important—lived experience.” Lived experience activated understanding and empathy, while clinical supervision and ongoing education provided strong facilitation skills and access to educational resources.

Our data also indicated that ongoing funding would not have been adequate without the respectful engagement that emerged from peer-empowered value constructs. The facilitator’s perceived support required more than financial support; Kenny also needed to feel that her expertise was valued, which in turn led to a level of investment and passion that was evident to participants and increased the sustainability of the program. Respondents repeatedly mentioned the facilitator’s passion for and investment in the program, which became a key part of the context for the next implementation phase. Refer to Table 4 for CMOCs 1.1-1.3 discussed in this section.

### Context 2: Accessible, sustainable program coordinated by a skilled peer facilitator

#### That is simply the magic of a skilled and passionate facilitator - PF Participant

Excavating the “magic” of Kenny’s approach presented one of the most difficult aspects of the analysis. We found that the combination of passion and skills she brought to the program was a key element of the second nested context: an accessible, sustainable program coordinated by a skilled peer facilitator that continued to rest on the foundation of strong organizational support. This helped bear the load of participant desperation that was compounded by the surrounding context of a lack of family support services amidst the intensity of the toxic drug crisis (Fernando et al., 2025; Hawkins, Salmon, et al., 2025). PF staff repeatedly mentioned the desperation exhibited by participants when they first joined the program, which was also reflected throughout participant narratives and helped explain the level of participant gratitude exhibited in our data.

The context of similar lived experience was also key. PF differentiated between the experiences of parents of adult children in addiction and the experiences of other affected family members, particularly parents of youth. Finding others with similar lived experience proved vital to participants’ acceptance of program resources.

We identified four main approaches that influenced participants’ responses to the program: 1) relational engagement; 2) non-pathological stance recognizing affected family members’ right to compassionate support; 3) balance of structure and adaptation; 4) orientation towards hope. Respondents spoke of Kenny’s relational approach, frequently mentioning her empathy and genuine caring: “It really comes across, her love for parents” (PF Participant). PF also exhibited a non-pathological approach towards addiction and also towards affected family members, insisting that family members needed and deserved support independent of their loved one’s trajectory. Participants also repeatedly mentioned Kenny’s facilitation skills in “moving things along,” following an agenda while still allowing participants time to share without allowing any one individual to “torpedo” the group. At the same time, the program’s framework included regular evaluations, providing a mechanism for feedback. This mix of structure and adaptation built trust and appreciation. Lastly, the group meetings consistently provided space to focus on positive progress, and Kenny frequently brought previous PF participants to the meetings to share encouraging stories. Teaching on “reasonable hope” (Weingarten, 2010) fostered positivity while still acknowledging hardship.

These underlying approaches engendered a sense of mutual connection, safety, welcome, trust, appreciation, and a willingness to engage in the program. A PF staff member reported that “the one thing that folks say when they leave the meeting for the first time is ‘I feel really accepted and understood and not blamed or judged. I found a home’” (PF Staff). When discussing the ability of PF participants to genuinely connect and be understood, one participant commented, “I mean just talking about this, I can physically feel the relief” (PF Participant). This led to the third nested context: a positive group culture with high levels of engagement with the support model. Refer to Table 4 for CMOCs 2.1-2.4 discussed in this section.

### Context 3: Strong, positive, engaged group culture supporting key program pillars

#### I’m making progress, slow but sure. — Evaluation Respondent

While our data indicated a strong group culture with a high level of program engagement, it was clear that not everyone engaged all the types of resources, nor to the same degree. Fully engaging the program required a willingness to hear others’ stories, some of which could be “horrific” or “raw.” Some participants did not join the program immediately after they initially encountered it because they “didn’t feel ready,” and others could not join because they found listening to others’ experiences too “heavy.” Participants who were ready to engage, however, describe multiple benefits in hearing others’ stories.

Parents Forever offered three main mechanisms of support: 1) peer support characterized by mutual understanding, non-judgement and compassion; 2) education and resources offered as a specific agenda item and occasional sessions with experts; 3) skill-building facilitated by peers and peer-developed resources as well as in one-on-one sessions with the program facilitator when needed. Skill-building primarily addressed differentiating what circumstances were under participants’ control, setting appropriate boundaries, and communication. It was clear that some participants attended primarily for emotional support, and others attended primarily for education or skill-development; however, many evaluation respondents and interview participants expressed deriving benefit from both.

Discrepancies in levels of engagement with educational and skill-building components of the program sometimes presented internal tension for participants. While they affirmed the expectation of compassion and non-judgement, interview participants and evaluation respondents mentioned a sense of unease with other participants who did not seem ready to apply the program’s learnings and seemed “stuck” in continual patterns of harm that were difficult to watch at times. These participants attested that sometimes they felt sad or frustrated when others in the group did not appear to move forward. This was reflected in the types of outcomes observed: while most reported improvements in personal wellbeing, not everyone reported interpersonal improvements with their loved one. Refer to Table 4 for CMOCs 3.1-3.3 discussed in this section.

### Simplified IPT

In line with our participatory approach, we reviewed our final CMOCs with Kenny and members from our research team with experience designing and implementing service programs. They felt that the CMOCs in the IPT were accurate, but the configurations were deemed too complex to be of much use guiding further implementation. They recommended something more visual that was simplified to describe the general outlines of the theory, expanding the CMO configuration to fit a broader program structure. Figure 2 displays a simplified IPT with more relevance to program partners who were interested in further implementation of PF’s model.

**Figure 2.**
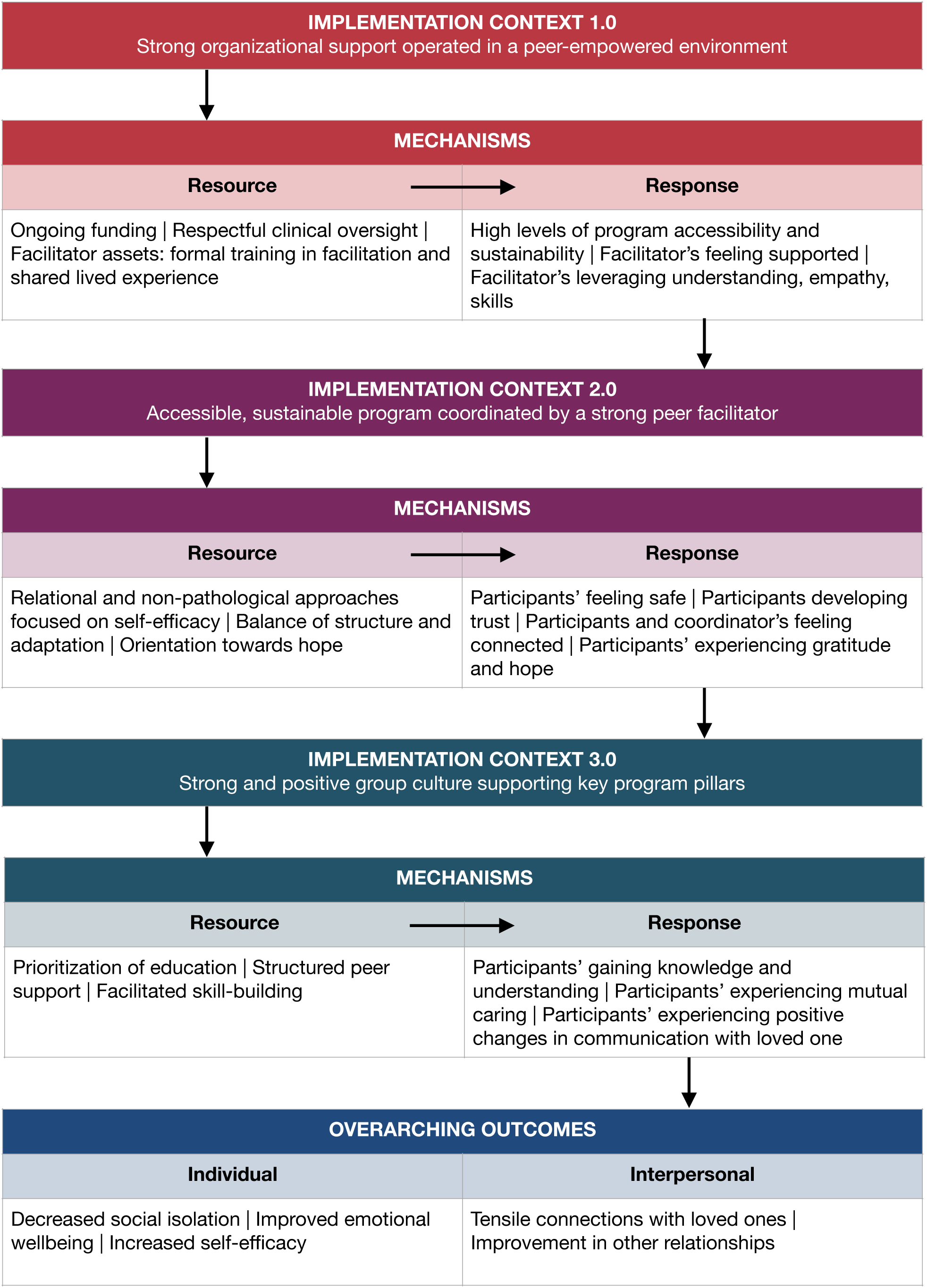
Simplified program theory in nested contexts

## Discussion

### Implications

Programs aligned with the SSCS model may provide a corrective approach in a service landscape dominated by programs heavily influenced by pathologization of affected family members and codependency theory (Denning, 2010; Devaney, 2017; Hawkins, Salmon, et al., 2025). The SSCS model asserts that affected family members attempt to cope under enormously stressful circumstances, and they require support in their own right that recognizes the challenges of their situation without blame (Orford, Velleman, et al., 2010). In contrast, codependency-derived frameworks may not provide adequate support in the context of the toxic drug crisis, particularly with their emphasis on cutting loved ones off rather than building skills to stay connected, which can in turn increase risks of harm in the context of a toxic drug crisis (Hawkins, Salmon, et al., 2025; Kelly et al., 2017). We found that family members responded to the program’s non-pathologizing stance towards their situations with trust, appreciation and a willingness to engage the program’s key pillars, ultimately leading to improvements in relationships with substance-using loved ones.

While codependency-derived programs still occupy a prominent place in North America, programs such as the Community Reinforcement Approach and Family Training (CRAFT), which are based on Social Network (SN) and Cognitive Behavioural Theory (CBT), have gained ground and have been shown to be effective (Assadbeigi et al., 2016). Nevertheless, these programs focus on improving the substance-using relative’s engagement with services rather than family member’s need for support in their own right (Roozen et al., 2010). Additionally, they often prioritize education and skill-building rather than providing emotional support.

However, we found that PF participants strongly attested to the importance of emotional support. Studies of peer support have shown that participants benefit from educational information (Peart et al., 2024), but they can be more effective when including both informational and emotional support (Kelly et al., 2017). The combination of emotional support, education and skill-building was a key distinctive of PF reflected in the SSCS framework.

Our study also reinforced the potential of third sector community groups to play a major role in social support that could fill gaps in formal services offered through health and social care systems (Morton et al., 2024). However, our IPT indicates that ongoing funding impacts longevity, which highlights the importance of examining financial sustainability. While a recent systematic review and meta-analysis demonstrate benefits in health and social services from peer support, few studies examine cost-effectiveness (Price et al., 2022). In the UK and Australia, studies report cost-savings from peer support, particularly group-based support; however, the evidence is mixed depending on the settings and methods, with many studies being of poor methodological quality (Bagnall et al., 2015; Wingate et al., 2017). With clear evidence of efficacy but less evidence of cost-benefit, additional research is needed that examines the cost-effectiveness of group peer support for affected family members.

### Limitations

Our IPT is based on a smaller-scale intervention that might not be as representative of other peer support groups. Additionally, we only recruited a small number of interview participants, although the diversity of our data addressed this limitation. Lastly, only one researcher conducted the initial coding, which was mitigated by our participatory methods and the retroductive analysis involving additional researchers.

## Conclusion

Our IPT suggests the following: 1) Strong organizational support in an environment that values lived expertise will provide a foundation of program sustainability by activating ongoing funding and the strengths of a peer facilitator; 2) A skilled facilitator with shared lived experience will leverage unique approaches to foster a positive group culture. These approaches involve relational interactions, a non-pathologized stance on substance use and affected families, structured support balanced with peer-driven adaptation, and an orientation towards hope; 3) A group culture characterized by safety, trust, genuine connection, gratitude and hope will activate the program’s pillars of emotional support, education and skill-building to produce individual and interpersonal outcomes. These outcomes include reduced social isolation, improved emotional wellbeing, increased self-efficacy, and improvements in relationships. Individual readiness has been shown to impact the level of engagement and outcomes in peer support (Garn et al., 2021). We likewise found that a readiness to hear others’ stories impacted program engagement at the outset, and a readiness to apply skill-building tools impacted whether or not participants received both individual and interpersonal benefits from participating.

We anticipate that further research will allow us to test and refine our IPT to support broader implementation of peer support groups in third sector settings that could prove to be cost-effective in addition to the positive health outcomes already documented in the literature. More research is needed into the cost-benefits of peer support groups and strategies for ensuring financial sustainability of these services over the long term.

## Disclosures

The authors report there are no competing interests to declare.

## Acknowledgements

The authors would like to thank the following individuals who are part of the core participatory action research team under which this study was conducted: Dr. Steven Esau, Marinel Kniseley, Connie Long, James Robson, Daniel Snyder and Mike Sikora.

## Data Availability

Data was collected and managed in concordance with the University of British Columbia Research and Providence Health Care Ethics Boards, which do not permit access to this data. Requests to access data for secondary analysis may be submitted to the corresponding author upon reasonable request.

## Funding Details

This work was supported by the Canadian Institute for Health Research, grant number F22-01474.

## Ethical Statement

Ethical approval for the research was granted by the University of British Columbia-Providence Health Care Research Ethics Board (H23-03688). All participants gave informed consent.

## Notes

### Competing Interest Statement

The authors have declared no competing interest.

### Author Declarations

Ethical approval for the research was granted by the University of British Columbia-Providence Health Care Research Ethics Board (H23-03688).

### Summary of Updates

Some of the content has been reorganized to include a Conclusion section, and the Abstract has also been reorganized to reduce the number of sub-sections. The authors added an additional researcher in the data analysis section to correct a previous omission. The style has been changed from Vancouver to APA.

## References

Assadbeigi, H., Pourshahbaz, A., Mohamadkhani, P., & Farhoudian, A. (2016). Effectiveness of Community Reinforcement and Family Training (CRAFT) on Quality of Life and Depression in Families with Drug Abuse. Global Journal of Health Science, 9(3), 167. 10.5539/gjhs.v9n3p167

Bagnall, A.-M., South, J., Hulme, C., Woodall, J., Vinall-Collier, K., Raine, G., Kinsella, K., Dixey, R., Harris, L., & Wrigh, N. M. (2015). A systematic review of the effectiveness and cost-effectiveness of peer educaiton and peer support in prisons. BMC Public Health, 15(290). 10.1186/s12889-015-1584-x

BC Coroner’s Service. (2023, November). BC Coroners Service death review panel: An urgent response to a continuing crisis. https://www2.gov.bc.ca/assets/gov/birth-adoption-death-marriage-and-divorce/deaths/coroners-service/death-review-panel/an_urgent_response_to_a_continuing_crisis_report.pdf

Belaid, L., Sarmiento, I., Dion, A., Pimentel, J. P., Rojas-Cárdenas, A., & Andersson, N. (2023). How does participatory research work: Protocol for a realist synthesis. BMJ Open, 13(9). 10.1136/bmjopen-2023-074075

Bellamy, C., Schmutte, T., & Davidson, L. (2017). An update on the growing evidence base for peer support. Mental Health and Social Inclusion, 21(3), 161–167. 10.1108/MHSI-03-2017-0014

Berg, R., Page, S., & Øgård-Repål. (2021). The effectiveness of peer-support for people living with HIV: A systematic review and meta-analysis. The Effectiveness of Peer-Support for People Living with HIV: A Systematic Review and Meta-Analysis. 10.1371/journal.pone.0252623

Copello, A., Templeton, L., Orford, J., & Velleman, R. (2010). The 5-Step Method: Principles and practice. Drugs: Education Prevention and Policy, 17, 86–99. 10.3109/09687637.2010.515186

Denning, P. (2010). Harm reduction therapy with families and friends of people with drug problems. Journal of Clinical Psychology, 66(2), 164–174. 10.1002/jclp.20671

Denomme, W. J., & Benhanoh, O. (2017). Helping concerned family members of individuals with substance use and concurrent disorders: An evaluation of a family member-oriented treatment program. Journal of Substance Abuse Treatment, 79, 34–45. 10.1016/j.jsat.2017.05.012

Devaney, E. (2017). The emergence of the affected adult family member in drug policy discourse: A Foucauldian perspective. *Drugs: Education*, Prevention and Policy, 24(4), 359–367. 10.1080/09687637.2017.1340433

Dopson, S., & Fitzgerald. (2007). Knowledge to action? Evidence-based health care in context. International Journal of Integrated Care, 7, e24.

Fernando, S., Hawkins, J., Kniseley, M., Sikora, M., Robson, J., Snyder, D., Battle, C., & Salmon, A. (2022). The Overdose Crisis and Using Alone: Perspectives of People Who Use Drugs in Rural and Semi-Urban Areas of British Columbia. Substance Use & Misuse, 57(12), 1864–1872. 10.1080/10826084.2022.2120361

Fernando, S., Hawkins, J., Salmon, A., Kniseley, M., Long, C., Esaue, S., Sikora, M., Snyder, D., & Robson, J. (2025). Stress and social support for substance affected carers in a toxic drug emergency. medRxiv. 10.1101/2025.10.20.25336204

Flint-Taylor, J., Abuhamdia, M., Berrrado, I., Bush, M., El Khoury, R., Fawzi, F., Jalal, M., Mourabiti, I., Sayah, H., Shukr, R., Wakim, N., Ward, B., & Stewart, S. (2023). Valuing adaptive programming: A study of resilience processes and outcomes. Evaluation and Program Planning, 98. 10.1016/j.evalprogplan.2023.102300

Flynn, R., Mrklas, K., Campbell, A., Wasylak, T., & Scott, S. D. (2021). Contextual factors and mechanisms that influence sustainability: A realist evaluation of two scaled, multi-component interventions. BMC Health Services Research, 21(1), 1194. 10.1186/s12913-021-07214-5

Garn, S. D., Glümer, C., Villadsen, S. F., Malling, G. M. H., & Christensen, U. (2021). Understanding the mechanisms generating outcomes in a Danish peer support intervention for socially vulnerable people with type 2-diabetes: A realist evaluation. Archives of Public Health, 79, 160. 10.1186/s13690-021-00676-3

Gilmore, B., McAuliffe, E., Power, J., & Vallières, F. (2019). Data Analysis and Synthesis Within a Realist Evaluation: Toward More Transparent Methodological Approaches. International Journal of Qualitative Methods. https://journals.sagepub.com/doi/full/10.1177/1609406919859754#bibr30-1609406919859754

Greenhalgh, T., Wong, G., Jagosh, J., Greenhalgh, J., Manzano, A., Westhorp, G., & Pawson, R. (2015). Protocol—the RAMESES II study: Developing guidance and reporting standards for realist evaluation. 10.1136/bmjopen-2015-008567

Haines, K. J., Beesley, S. J., Hopkins, R. O., McPeake, J., Quasim, T., Ritchie, K., & Iwashyna, T. J. (2018). Peer Support in Critical Care: A Systematic Review. Critical Care Medicine, 46(9), 1522. 10.1097/CCM.0000000000003293

Halsall, T., Daley, M., Hawke, L., Henderson, J., & Matheson, K. (2022). “You can kind of just feel the power behind what someone’s saying”: A participatory-realist evaluation of peer support for young people coping with complex mental health and substance use challenges. BMC Health Services Research, 22. 10.1186/s12913-022-08743-3

Hawkins, J., Kniseley, M., Fernando, S., Salmon, A., Robson, J., Sikora, M., & Esau, S. (2025). Breaking Bread, Building Hope: An Eight-Year Partnership Addressing the Toxic Drug Crisis in British Columbia. SocArXiv. 10.31235/osf.io/q794j_v1

Hawkins, J., Salmon, A., Fernando, S., Battle, C., Esau, S., Snyder, D., & Sikora, M. (2025). ‘I don’t know what we should have done differently’: A qualitative study on the dilemmas of ‘tough love’ and toxic drugs in British Columbia, Canada. Drugs: Education Prevention and Policy, 1–10. 10.1080/09687637.2025.2493140

Jagosh, J. (2019). Realist Synthesis for Public Health: Building an Ontologically Deep Understanding of How Programs Work, For Whom, and In Which Contexts. Annual Review of Public Health, 40, 361–372. 10.1146/annurev-publhealth-031816-044451

Jagosh, J., Bush, P. L., Salsberg, J., Macaulay, A. C., Greenhalgh, T., Wong, G., Cargo, M., Green, L. W., Herbert, C. P., & Pluye, P. (2015). A realist evaluation of community-based participatory research: Partnership synergy, trust building and related ripple effects. BMC Public Health, 15(1), 725. 10.1186/s12889-015-1949-1

Jagosh, J., Macaulay, A. C., Pluye, P., Salsberg, J., Bush, P. L., Henderson, J., Sirett, E., Wong, G., Cargo, M., Herbert, C. P., Seifer, S. D., Green, L. W., & Greenhalgh, T. (2012). Uncovering the Benefits of Participatory Research: Implications of a Realist Review for Health Research and Practice. The Milbank Quarterly, 90(2), 311–346. 10.1111/j.1468-0009.2012.00665.x

Jagosh, J., Stott, H., Halls, S., Thomas, R., Liddiard, C., Cupples, M., Cramp, F., Kersten, P., Foster, D., & Walsh, N. E. (2022). Benefits of realist evaluation for rapidly changing health service delivery. BMJ Open, 12(7). 10.1136/bmjopen-2021-060347

Kelly, J. F., Fallah-Sohy, N., Cristello, J., & Bergman, B. (2017). Coping with the enduring unpredictability of opioid addiction: An investigation of a novel family-focused peer-support organization. Journal of Substance Abuse Treatment, 77, 193–200. 10.1016/j.jsat.2017.02.010

Kundurthi, V., Reddy, S. K., Jagannathan, A., Berigai Parthasaratyhy, N., & L, P. (2025). Effectiveness of Peer Support Group Interventions for Persons with Mental Illness: A Systematic Review. Journal of Psychosocial Rehabilitation and Mental Health. 10.1007/s40737-025-00486-8

Marchal, B., van Belle, S., van Olmen, J., Hoerée, T., & Kegels, G. (2012). Is realist evaluation keeping its promise? A review of published empirical studies in the field of health systems research. Evaluation, 18(2), 192–212. 10.1177/1356389012442444

Mathias, H., Auger, S., Schulz, P., & Hyshka, E. (2025). Including Families in a Response to the Unregulated Toxic Drug Crisis: A Call to Action. Substance Use & Misuse, 60(3), 452–456. 10.1080/10826084.2024.2431042

Mathias, H., Duff, E., Schulz, P., Auger, S., Gravel-Ouellette, A., Lockhart, T., McCorriston, W., McCrindle, J., Mirza, N., Pijl, E., Savard, T., & Hyshka, E. (2025). Rural community-based participatory research with families of people who use drugs: Key considerations from a multi-provincial research partnership. Harm Reduction Journal, 22(1). 10.1186/s12954-025-01247-3

Morton, T., Evans, S. B., Swift, R., Bray, J., Frost, F., Russell, C., Brooker, D., Wong, G., & Hullah, N. (2024). Strategic and operational issues in sustaining community-based dementia support groups: The Get Real with Meeting Centres realist evaluation part 2. Aging & Mental Health, 28(12), 1704–1712. 10.1080/13607863.2024.2372058

Natung, B., Awuor, E., Mall, M., & Mishra, M. (2025). The role of self-help groups in social capital development: A systematic review. Cogent Social Sciences, 11(1), 2527393. 10.1080/23311886.2025.2527393

Orford, J., Copello, A., Velleman, R., & Templeton, L. (2010). Family members affected by a close relative’s addiction: The stress-strain-coping-support model. *Drugs: Education*, Prevention and Policy, 17(sup1), 36–43. 10.3109/09687637.2010.514801

Orford, J., Velleman, R., Copello, A., Templeton, L., & Ibanga, A. (2010). The experiences of affected family members: A summary of two decades of qualitative research. Drugs: Education Prevention and Policy, 17, 44–62. 10.3109/09687637.2010.514192

Orford, J., Velleman, R., Natera, G., Templeton, L., & Copello, A. (2013). Addiction in the family is a major but neglected contributor to the global burden of adult ill-health. Social Science & Medicine (1982), 78, 70–77. 10.1016/j.socscimed.2012.11.036

Pawson, R., & Tilley, N. (1997). Realistic Evaluation. Sage Publications. https://uk.sagepub.com/en-gb/eur/realistic-evaluation/book205276

Peart, A., Horn, F., Grigg, J., Manning, V., Campbell, R., & Lubman, D. I. (2023). Online Peer-Led Support Program for Affected Family Members of People Living with Addiction: A Mixed Methods Study. International Journal of Mental Health and Addiction. 10.1007/s11469-023-01082-2

Peart, A., Horn, F., Manning, V., Campbell, R., & Lubman, D. I. (2024). The experiences of family members attending an online addiction education program: A qualitative study. *Drugs: Education*, Prevention and Policy, 31(3), 310–317. 10.1080/09687637.2023.2184248

Pfeiffer, P. N., Heisler, M., Piette, J. D., Rogers, M. A. M., & Valenstein, M. (2011). Efficacy of peer support interventions for depression: A meta-analysis. General Hospital Psychiatry, 33(1), 29–36. 10.1016/j.genhosppsych.2010.10.002

Price, A., de Bell, S., Shaw, N., Bethel, A., Anderson, R., & Coon, J. T. (2022). What is the volume, diversity and nature of recent, robust evidence for the use of peer support in health and social care? An evidence and gap map. Campbell Systematic Reviews, 18(3). 10.1002/cl2.1264.

Rokiyah, R., Zuanda, N., Alrefi, A., & Akbari, A. (2024). The Role of Social Support in Substance Addiction Recovery: A Systematic Review. Edusoshum : Journal of Islamic Education and Social Humanities, 4(2), 213–222. 10.52366/edusoshum.v4i2.112

Roozen, H. G., De Waart, R., & Van Der Kroft, P. (2010). Community reinforcement and family training: An effective option to engage treatment-resistant substance-abusing individuals in treatment. Addiction, 105(10), 1729–1738. 10.1111/j.1360-0443.2010.03016.x

Salmon, A., Hawkins, J., & Kenny, F. (2025, May 23). Supporting affected family members in the Fraser East of British Columbia: Reflections on needs, theories, peer support, and learnings from a realist evaluation. AFINet 2025 Conference. Addiction, Family Members, and Affected Others International Conference, 6th Edition, Quebec City.

Sarkies, M. N., Francis-Auton, E., Long, J. C., Pomare, C., Hardwick, R., & Braithwaite, J. (2022). Making implementation science more real. BMC Medical Research Methodology, 22, 178. 10.1186/s12874-022-01661-2

Selbekk, A. S., Adams, P. J., & Sagvaag, H. (2018). “A Problem Like This Is Not Owned by an Individual”: Affected Family Members Negotiating Positions in Alcohol and Other Drug Treatment. Contemporary Drug Problems, 45(2), 146–162. 10.1177/0091450918773097

Selbekk, A. S., & Sagvaag, H. (2016). Troubled families and individualised solutions: An institutional discourse analysis of alcohol and drug treatment practices involving affected others. Sociology of Health & Illness, 38(7), 1058–1073. 10.1111/1467-9566.12432

Shorey, S., & Chua, J. Y. X. (2022). Effectiveness of peer support interventions for adults with depressive symptoms: A systematic review and meta-analysis. Journal of Mental Health, 32(2), 465–479. 10.1080/09638237.2021.2022630

Slaunwhite, A. K., Ronis, S. T., Sun, Y., & Peters, P. A. (2017). The emotional health and well-being of Canadians who care for persons with mental health or addictions problems. Health & Social Care in the Community, 25(3), 840–847. 10.1111/hsc.12366

Soares, A., Ferreira, G., & Pereira, M. G. (2016). Depression, distress, burden and social support in caregivers of active versus abstinent addicts. Addiction Research and Theory, 24. 10.3109/16066359.2016.1173681

Wallerstein, N., & Duran, B. (2010). Community-Based Participatory Research Contributions to Intervention Research: The Intersection of Science and Practice to Improve Health Equity. American Journal of Public Health, 100(Suppl 1), S40–S46. 10.2105/AJPH.2009.184036

Weingarten, K. (2010). Reasonable hope: Construct, clinical applications, and supports. Family Process, 49(1), 5–25. 10.1111/j.1545-5300.2010.01305.x

White, S., Foster, R., Marks, J., Morshead, R., Goldsmith, L., Barlow, S., Sin, J., & Gillard, S. (2020). The effectiveness of one-to-one peer support in mental health services: A systematic review and meta-analysis | BMC Psychiatry. BMC Psychiatry, 20. https://link.springer.com/article/10.1186/s12888-020-02923-3

Wingate, L., Graffy, J., Holman, D., & Simmons, D. (2017). Can peer support be cost saving? An economic evaluation of RAPSID: a randomized controlled trial of peer support in diabetes compared to usual care alone in East of England communities. BMJ Open Diabetes Research & Care, 5(1). https://drc.bmj.com/content/5/1/e000328

Wong, G., Westhorp, G., Manzano, A., Greenhalgh, J., Jagosh, J., & Greenhalgh, T. (2016). RAMESES II reporting standards for realist evaluations. BMC Medicine, 14(1), 96. 10.1186/s12916-016-0643-1

Wong, J., Delormier, T. W., Bergeron, D., Chan, H. M., Gabriel-Ferland, P., & Wenniserí:iostha, J. (2025). “If you show them respect, you’re going to [get] respect back”: A theory for engaging First Nations for knowledge translation within a national nutrition and health survey. BMC Public Health, 25. 10.1186/s12889-025-22196-3

